# Prognostic value of advanced lung cancer inflammation index (ALI) combined with geriatric nutritional risk index (GNRI) in patients with chronic heart failure

**DOI:** 10.1101/2023.07.07.23292398

**Authors:** Tao Shi, Yan Wang, Yunzhu Peng, Meifen Wang, Yanji Zhou, Wenyi Gu, Yanyan Li, Jie Zou, Na Zhu, Lixing Chen

## Abstract

**Background:** This study was undertaken to explore the predictive value of the advanced lung cancer inflammation index (ALI) combined with the geriatric nutritional risk index (GNRI) for all‐cause mortality in patients with CHF.

**Methods and Results:** We enrolled 1123 patients with HF admitted to our cardiology department from January 2017 to October 2021. Patients were divided into Group 1 (ALI<24.60 and GNRI<94.41), Group 2 (ALI<24.60 and GNRI≥94.41), Group 3 (ALI≥24.60 and GNRI<94.41) and Group 4 (ALI≥24.60 and GNRI≥94.41), according to the median ALI and GNRI. From the analysis of the relationship between the ALI and GNRI, we concluded that there was a mild positive linear correlation (r= 0.348, p< 0.001) and no interaction (p=0.140) between the ALI and GNRI. Kaplan‒Meier analysis showed that the cumulative incidence of all‐cause mortality in patients with CHF was highest in Group 1 (log‐rank χ^2^ 126.244, p<0.001). Multivariate Cox proportional hazards analysis revealed that ALI and GNRI were independent predictors of all‐cause mortality in CHF patients (ALI: HR 0.313, 95% CI 0.228‐0.430, p <0.001; GNRI: HR 0.966, 95% CI 0.953‐0.979, p <0.001). The area under the curve (AUC) for ALI combined with GNRI was 0.711 (p<0.001), according to the time‐dependent ROC curve.

**Conclusion:** ALI and GNRI were independent predictors of all‐cause mortality in CHF patients. Patients with CHF had the highest risk of all‐cause mortality when the ALI was <24.60 and the GNRI was <94.41. ALI combined with the GNRI has good predictive value for the prognosis of CHF patients.

## Introduction

Heart failure (HF) is the major cause of morbidity and mortality worldwide and is on the increase^1^. China is the most populous country in the world, so the total number of HF patients is enormous. The incidence and cost of treatment for chronic HF (CHF) patients are likely to increase in the next decade due to the ageing population^2^. HF imposes a huge financial burden on patients and is a significant resource drain on health care systems, and HF is becoming a global problem^3^.

Patients with CHF are characterized by high levels of inflammation, hypercatabolic syndrome with weight loss, skeletal sarcopenia and muscle wasting, and clinically, these patients are often severely malnourished because nutritional interventions are often lacking or neglected^4^. Malnutrition in CHF patients sometimes progresses to overt “cardiac cachexia”, characterized by protein‐calorie malnutrition with muscle atrophy and peripheral oedemaedema^5^. Although such extreme states may be apparent, recognizing less severe malnutrition can be more difficult. It is therefore necessary to carry out a routine assessment of the nutritional status of patients with HF.

Several objective nutritional indices are often used in the HF field, among which the geriatric nutritional risk index (GNRI), controlling nutritional status (CONUT) score and prognostic nutrition index (PNI) are recognized prognostic markers in patients with HF^6–8^. Among these, the GNRI appears to be a relatively optimal tool for screening malnutrition and showed the highest prognostic value in outpatients with HF^6^. The advanced lung cancer inflammation index (ALI) is an emerging inflammatory and nutritional biomarker that includes NLR, BMI, and serum albumin. Several studies have reported the predictive value of the ALI in the prognosis of HF patients^9,10^. However, no studies have confirmed the predictive value of the ALI combined with the GNRI for the prognosis of patients with HF. Therefore, the aim of this study was to investigate the prognostic impact of the ALI combined with the GNRI in HF patients.

## Methods

### 1. Study population

We enrolled 1221 patients with a diagnosis of chronic heart failure (CHF) who were admitted to the First Affiliated Hospital of Kunming Medical University from January 2017 to October 2021. We included patients admitted with CHF classified as New York Heart Association (NYHA) functional class III or IV and a brain natriuretic peptide (BNP) level of ≥500 pg/mL. We included 1123 patients with CHF in this study after excluding those who lacked the necessary data (e.g., routine blood tests, cardiac ultrasound data or body mass index), died during hospitalization, had a combination of other serious diseases (e.g., malignancies, infectious diseases, blood diseases, or severe renal or liver dysfunction) or were lost to follow‐up.

### 2. Data collection

At the time of admission, demographic and clinical information, electrocardiograms, cardiac ultrasound data and blood samples were collected. Data included patient age and sex, heart rate (HR), blood pressure (BP), body mass index (BMI), New York Heart Association cardiac function classification (NYHA), medical history, brain natriuretic peptide (BNP), white blood cells (WBC), red blood cells (RBC), neutrophils (NBC), lymphocytes (LBC), haemoglobin (Hb), platelets (PLT), sodium, potassium, chlorine, albumin (Alb), alanine aminotransferase (ALT), aspartate aminotransferase (AST), creatinine (Cre), uric acid (UA), total cholesterol (TC) and estimated glomerular filtration rate (eGFR), QRS width, left atrial diameter (LAd), left ventricular end‐diastolic diameter (LVDd), right atrial diameter (RAd), right ventricular diameter (RVd), and left ventricular ejection fraction (LVEF). Blood samples were taken from all patients after an overnight fast (8‐12 hours) and sent to the laboratory of the First Affiliated Hospital of Kunming Medical University.

The following formula was used to derive the advanced lung cancer inflammation index (ALI): ALI = BMI × Alb/NLR, where BMI = weight in kilograms/(height in metres)^2^, Alb = serum albumin in grams per decilitre, and NLR (neutrophil‐to‐lymphocyte ratio) = absolute neutrophil count/absolute lymphocyte count^11^. The geriatric nutritional risk index (GNRI) was calculated according to the following formula: GNRI = 14.89 × Alb (g/dL) + 41.7 × BMI/22, where BMI/22 was set to 1 if the patient’s BMI/22 was greater than 1^12^.

The researchers collected the survival data by conducting telephone interviews with the patients or their family members. All‐cause mortality was the primary endpoint of the study.

### 3. Ethics

The study was approved by the Medical Ethics Committee of the First Affiliated Hospital of Kunming Medical University, and the study complied with the Declaration of Helsinki. Written informed consent was obtained from each patient in the study. The ethics number of the study was (2022) Ethics L No.173.

### 4. Statistical analysis

Patients with HF were divided into four groups according to the median ALI and GNRI: Group 1 (ALI<24.60 and GNRI<94.41), Group 2 (ALI<24.60 and GNRI≥94.41), Group 3 (ALI≥24.60 and GNRI<94.41) and Group 4 (ALI≥24.60 and GNRI≥94.41). When describing baseline patient characteristics, continuous variables are expressed as the mean ± standard deviation when normally distributed, otherwise as the median with interquartile range, while categorical variables are expressed as numbers and percentages. ALI and BNP were logarithmically transformed due to the highly skewed distribution of the original values. When comparing baseline characteristics between the four groups, variance analyses were used for normally distributed continuous variables, Mann‒Whitney U tests for nonnormally distributed data, and chi‐square tests for categorical variables. Correlation analysis based on Spearman’s nonparametric test and two‐way ANOVA interaction analysis were used to assess the correlation and interaction between the ALI and GNRI, respectively. To assess the relationship between the ALI and GNRI and all‐cause mortality, we used Kaplan‒Meier curves for analysis and log‐rank tests for comparison. Univariate Cox proportional hazard regression analyses were used to roughly show the effect of each variable on all‐cause mortality. Subsequently, multivariate Cox proportional hazard regression analysis was performed on variables with p values less than 0.05 in the univariate analysis to identify independent predictors of all‐cause mortality in patients with HF. Receiver operating characteristic (ROC) analysis was used to assess the predictive value of the ALI combined with the GNRI on the risk of all‐cause mortality in patients with CHF. This study used IBM SPSS Statistics version 26.0 and MedCalc for data analysis. A p value less than 0.05 was regarded as statistically significant.

## Results

### 1. Baseline patient characteristics

In this study, we included 1123 patients with CHF. Based on the median ALI and GNRI, we divided the patients into four groups: Group 1 (ALI<24.60 and GNRI<94.41, n=336), Group 2 (ALI<24.60 and GNRI≥94.41, n=225), Group 3 (ALI≥24.60 and GNRI<94.41, n=221) and Group 4 (ALI≥24.60 and GNRI≥94.41, n=341). There were statistically significant differences between the four groups in age, diastolic BP, BMI, NYHA, coronary disease, lg BNP, WBC, RBC, NBC, LBC, NLR, Hb, sodium, potassium, chlorine, Alb, Cre, UA, TC, eGFR, QRS wave, LAd, LVDd, and LVEF (p<0.05) (Table 1).

**Table 1.**
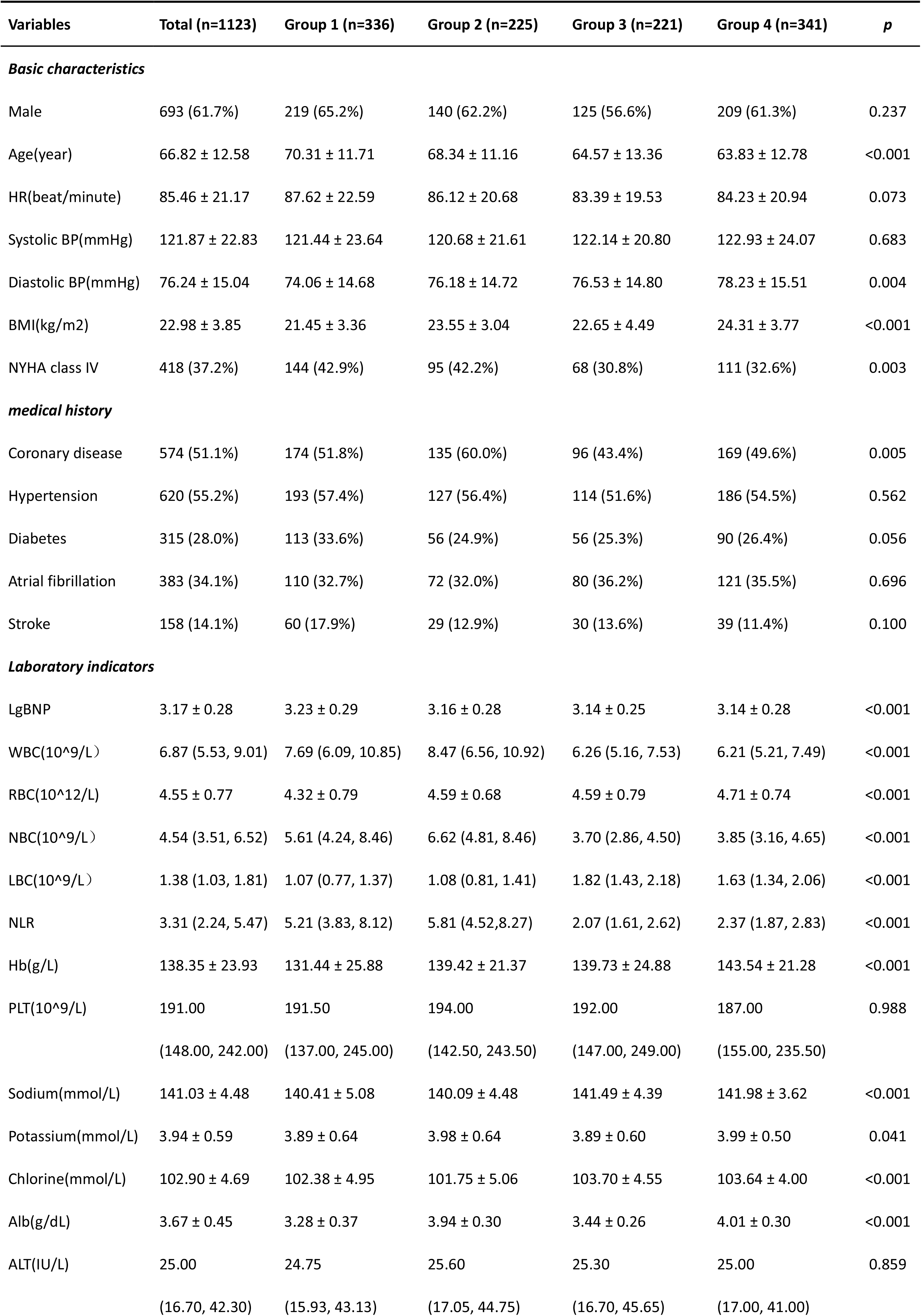
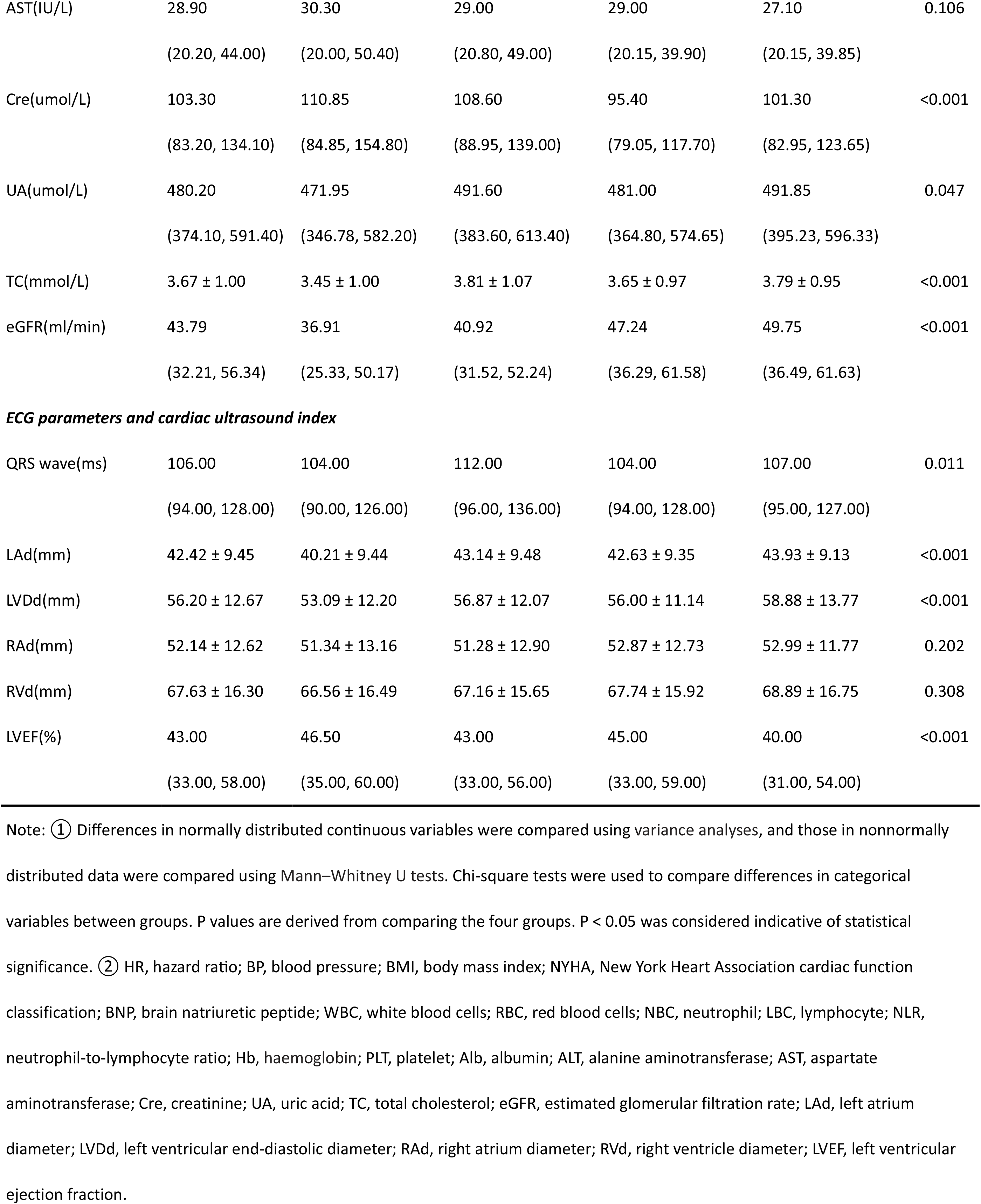
Baseline characteristics according to ALI combined with GNRI.

### 2. Correlation between advanced lung cancer inflammation index (ALI) and geriatric nutritional risk index (GNRI)

The GNRI is widely used in nutritional assessment, and the ALI is a novel inflammatory and nutritional biomarker. Therefore, we examined the correlation of the ALI and GNRI in CHF patients. A correlation analysis based on Spearman’s nonparametric test showed that the ALI was mildly positively linearly correlated with the GNRI in the total study population. [Spearman’s correlation coefficient (r): 0.348, p< 0.001] (Figure 1). Two‐way ANOVA revealed no interaction between the ALI and GNRI (p=0.140) (Figure 2).

**Figure 1.**
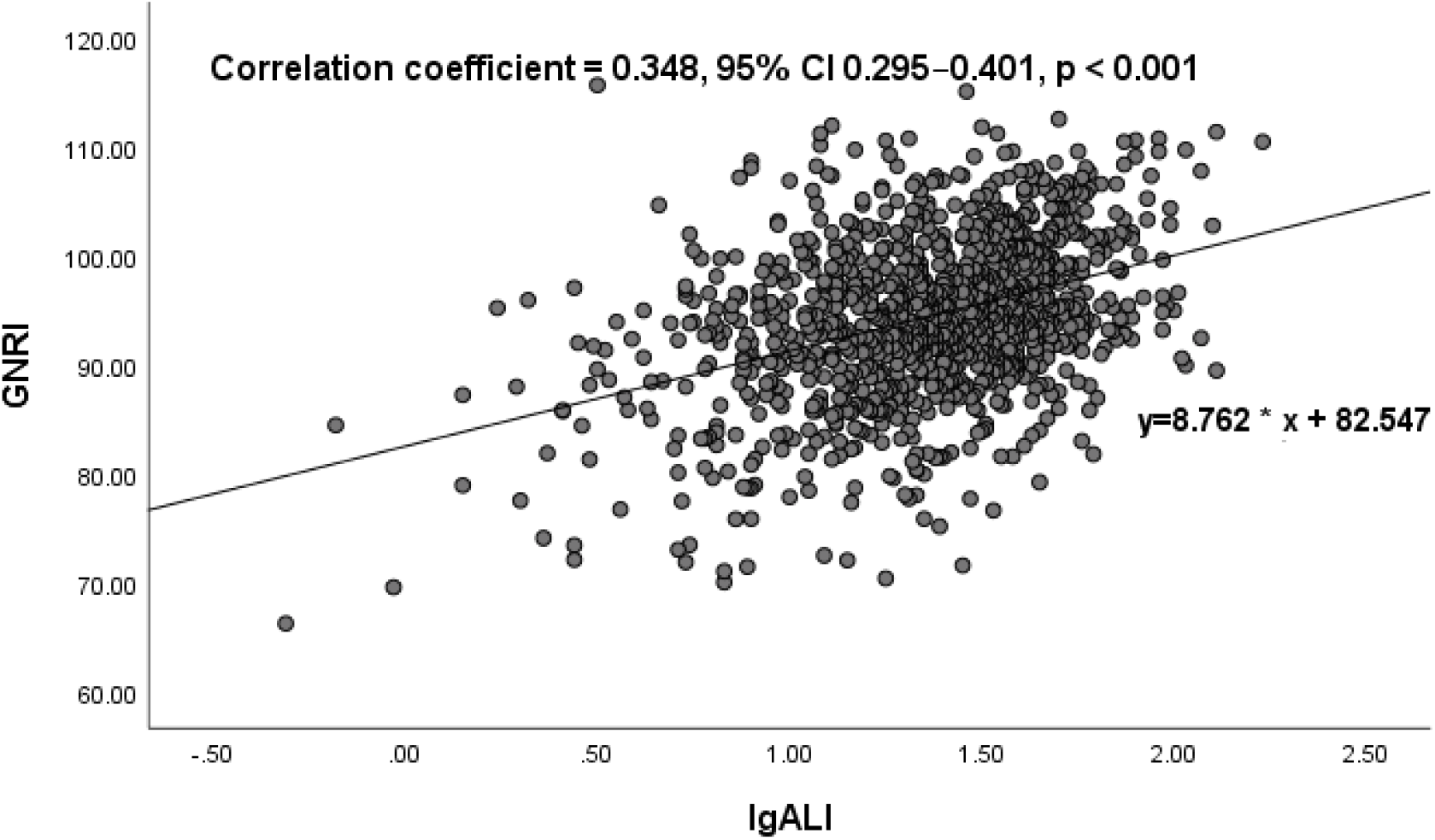
Correlation between advanced lung cancer inflammation index (ALI) and geriatric nutritional risk index (GNRI). The ALI was mildly positively linearly correlated with the GNRI (correlation coefficient = 0.348, 95% confidential interval (CI) 0.295–0.401, p < 0.001).

**Figure 2.**
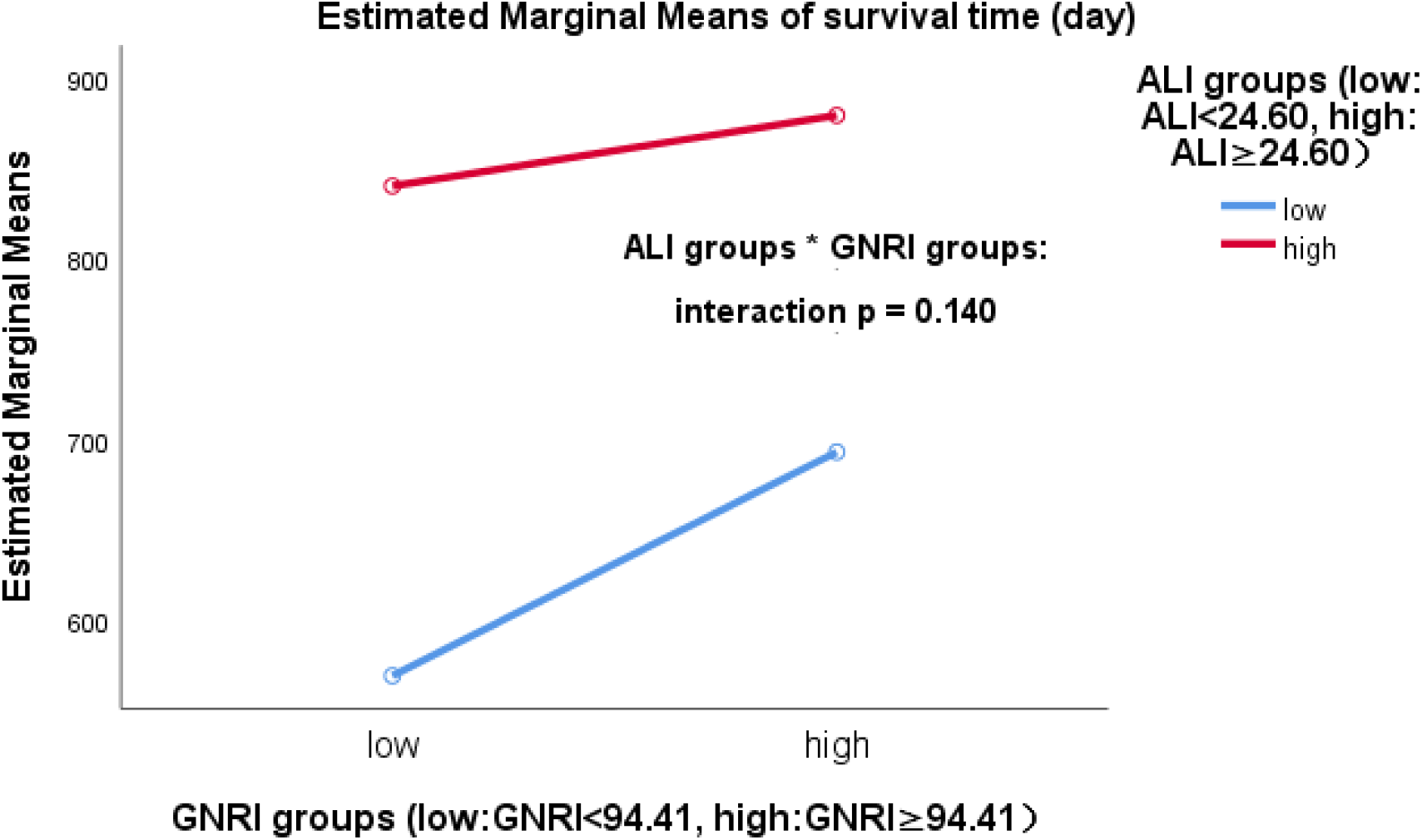
Interaction between advanced lung cancer inflammation index (ALI) and geriatric nutritional risk index (GNRI). There was no interaction between the ALI and GNRI (interaction p = 0.140).

### 3. Between advanced lung cancer inflammation index (ALI) and geriatric nutritional risk index (GNRI) and all‐cause mortality

To test the prognostic significance of the ALI combined with GMRI in patients with CHF, we conducted a Kaplan‒Meier analysis. The Kaplan‒Meier analysis showed that the cumulative incidence of all‐cause death was significantly higher in CHF patients in the low group than in those in the high group, either in the ALI group or the GNRI group (ALI: log‐rank χ^2^ 105.785, p<0.001; GNRI: log‐rank χ^2^ 27.731, p<0.001) (Figure 3‐4). When the ALI was combined with GNRI, the Kaplan‒Meier analysis revealed that the cumulative incidence of all‐cause mortality in CHF patients was highest in Group 1, second highest in Group 2 and tied for lowest in Group 3 and Group 4 (log‐rank χ^2^ 126.244, p<0.001) (Figure 5).

**Figure 3.**
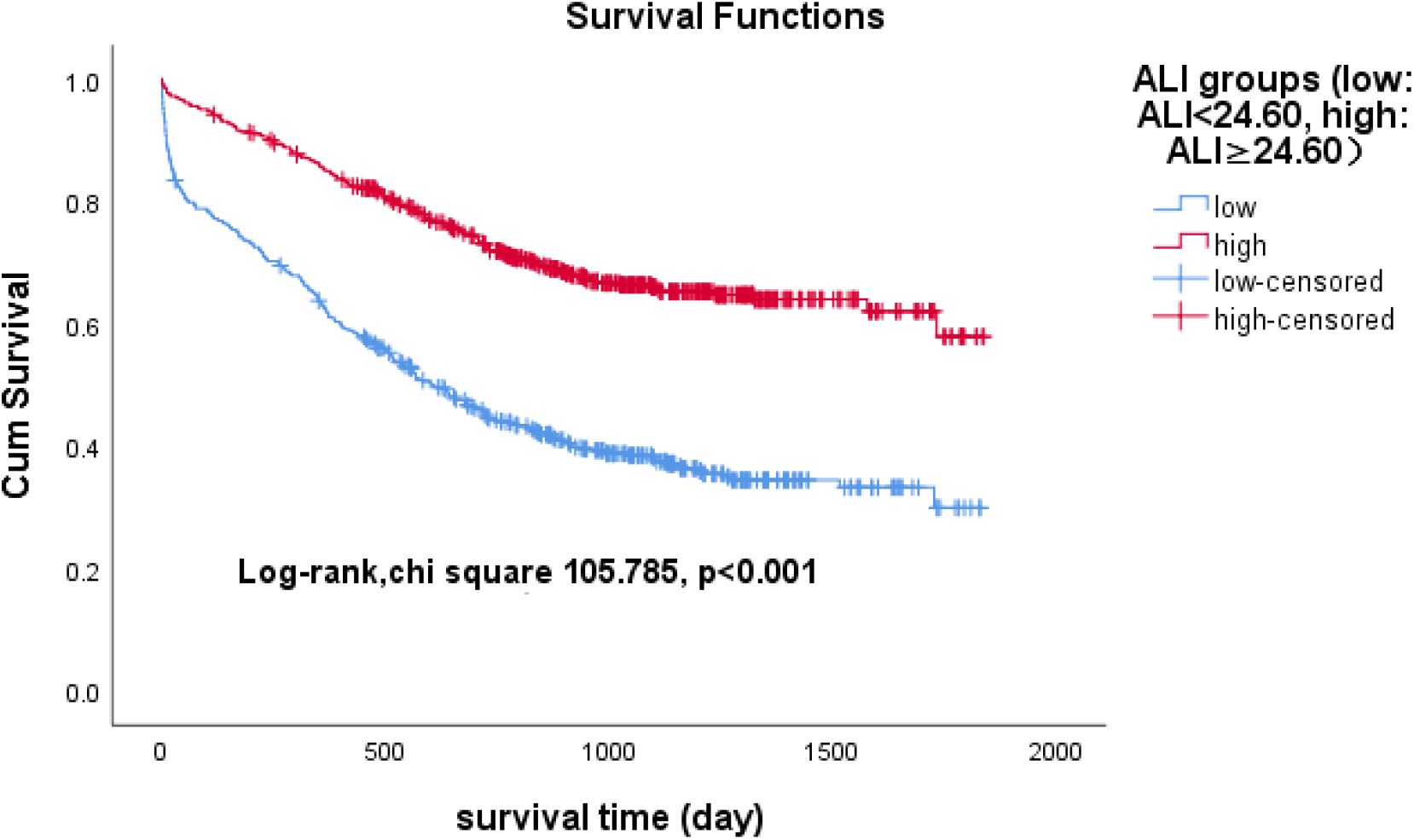
Kaplan‒Meier survival curves for CHF patients across the advanced lung cancer inflammation index (ALI).

**Figure 4.**
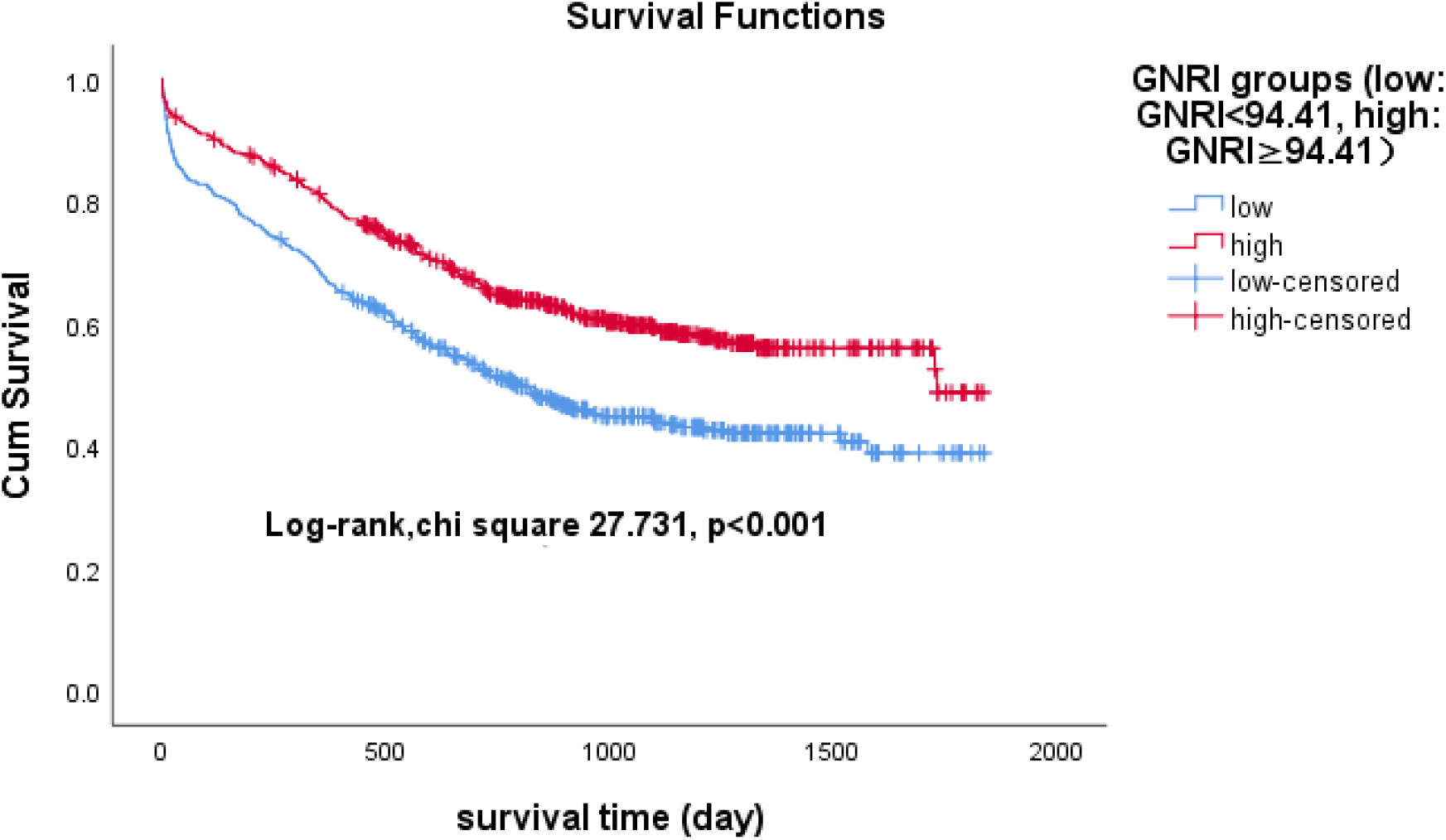
Kaplan‒Meier survival curves for CHF patients across the geriatric nutritional risk index (GNRI).

**Figure 5.**
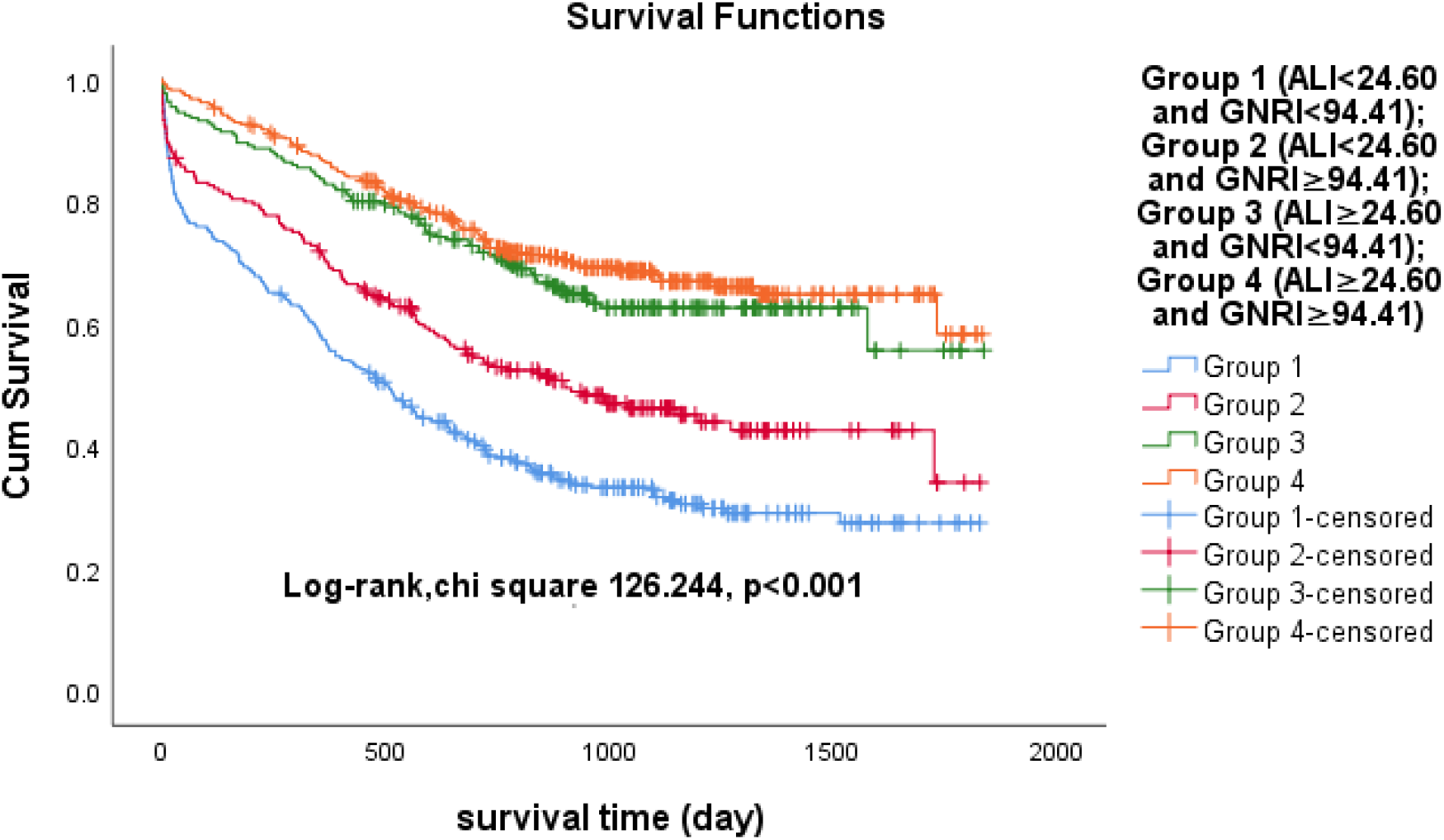
Kaplan‒Meier survival curves for CHF patients in the four groups. Group 1 (ALI<24.60 and GNRI<94.41); Group 2 (ALI<24.60 and GNRI≥94.41); Group 3 (ALI≥24.60 and GNRI<94.41); Group 4 (ALI≥24.60 and GNRI≥94.41).

### 4. Advanced lung cancer inflammation index (ALI) and geriatric nutritional risk index (GNRI) as predictors of adverse outcomes

After correcting for age, NYHA, heart rate, systolic BP, diastolic BP, ALT, AST, creatinine, uric acid, eGFR, sodium, chloride, and lg BNP, the multivariate Cox proportional hazards analyses showed that the ALI and GNRI were independent predictors of all‐cause death in patients with CHF. (ALI: HR 0.313, 95% CI 0.228‐0.430, p <0.001; GNRI: HR 0.966, 95% CI 0.953‐0.979, p <0.001) (Table 2). Hazard ratios for mortality in patients with CHF based on the median ALI and GNRI were calculated. The results show that, using Group 4 as a reference, the risk of death is 3.113 times higher for Group 1 (p<0.001), 2.067 times higher for Group 2 (p<0.001) and no different for Group 3 than for Group 4 (p=0.307) (Table 3). We can conclude that the risk of death in patients with CHF is highest when the ALI is <24.60 and GNRI <94.41.

**Table 2.**
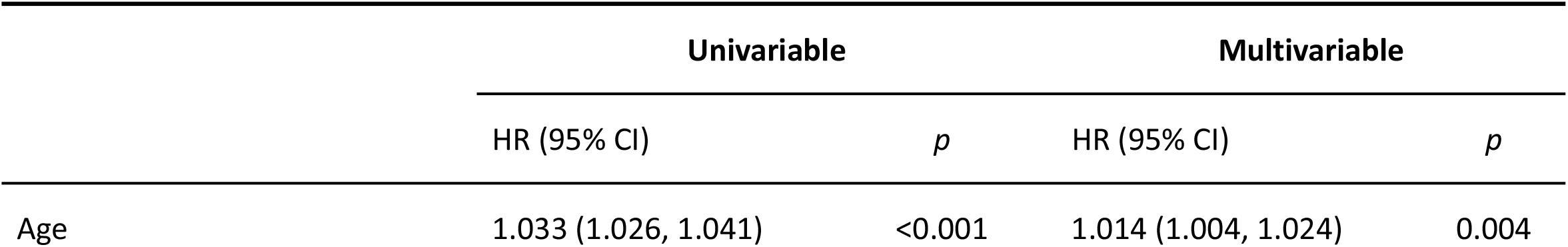
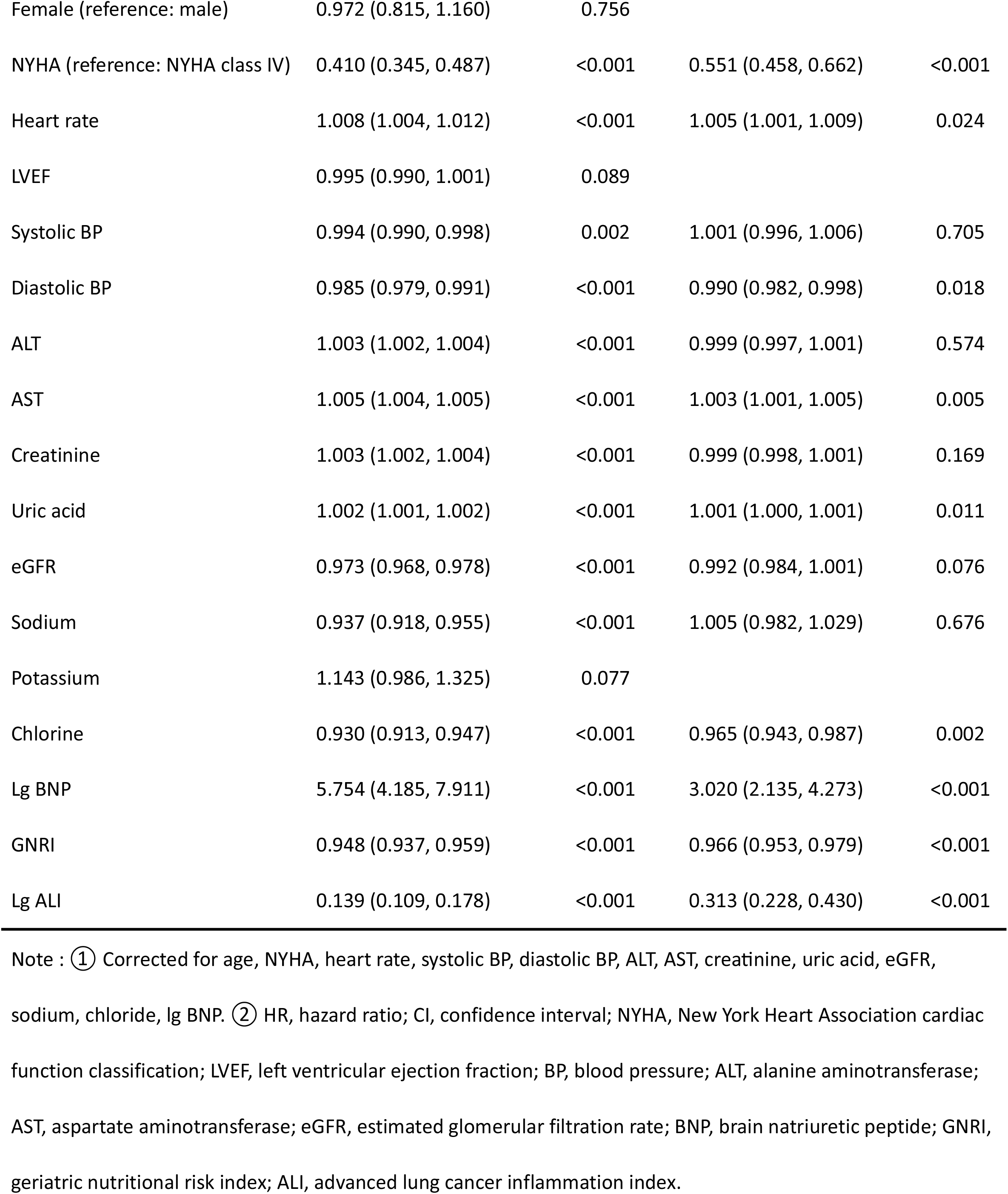
Results of univariable analysis and final model of multivariable analysis using Cox proportional hazard analysis of all‐cause.

**Table 3.** Hazard ratios for mortality of patients with CHF according to the medians of ALI and GNRI.

| Four Groups | ALI | GNRI | Mortality,% | HR (95% CI) | *p* |
| --- | --- | --- | --- | --- | --- |
| Group 4 (n=264) | ≥24.60 | ≥94.41 | 20.0% | Reference group |  |
| Group 1 (n=264) | <24.60 | <94.41 | 43.0% | 3.113 (2.468, 3.926) | p<0.001 |
| Group 2 (n=177) | <24.60 | ≥94.41 | 22.4% | 2.067 (1.589, 2.689) | p<0.001 |
| Group 3 (n=177) | ≥24.60 | <94.41 | 14.6% | 1.166 (0.869,1.564) | p=0.307 |

Note : ①Group 1, Group 2 and Group 3 all used Group 4 as the reference group. ②Group 1 (ALI<24.60 and GNRI<94.41); Group 2 (ALI<24.60 and GNRI≥94.41); Group 3 (ALI≥24.60 and GNRI<94.41); Group 4 (ALI≥24.60 and GNRI≥94.41).

### 5. Predictive ability of the advanced lung cancer inflammation index (ALI) combined with the geriatric nutritional risk index (GNRI) in patients with CHF

The time‐dependent ROC curves showed that the area under the curve (AUC) was 0.633 for GNRI (p<0.001), 0.704 for ALI (p<0.001) and 0.711 for ALI combined with GNRI (p<0.001) (Figure 6). To further investigate the predictive impact of the ALI combined with the GNRI, we built a basic Model (P1) that included age, NYHA, heart rate, systolic BP, diastolic BP, ALT, AST, creatinine, uric acid, eGFR, sodium, chloride, and lg BNP. We then added ALI and GNRI to the basic Model (P1) to form a new Model (P2). According to the time‐dependent ROC curves, the AUC for P1 was 0.785 with a sensitivity of 71.72% and a specificity of 75.55% (p<0.001); the AUC for P2 was 0.815 with a sensitivity of 79.39% and a specificity of 70.34% (p<0.001); and the AUC for P1 was greater than the AUC for P2 (P<0.001) (Figure 7). Based on the above analyses, we can conclude that the ALI combined with the GNRI had good predictive power for the prognosis of CHF patients.

**Figure 6.**
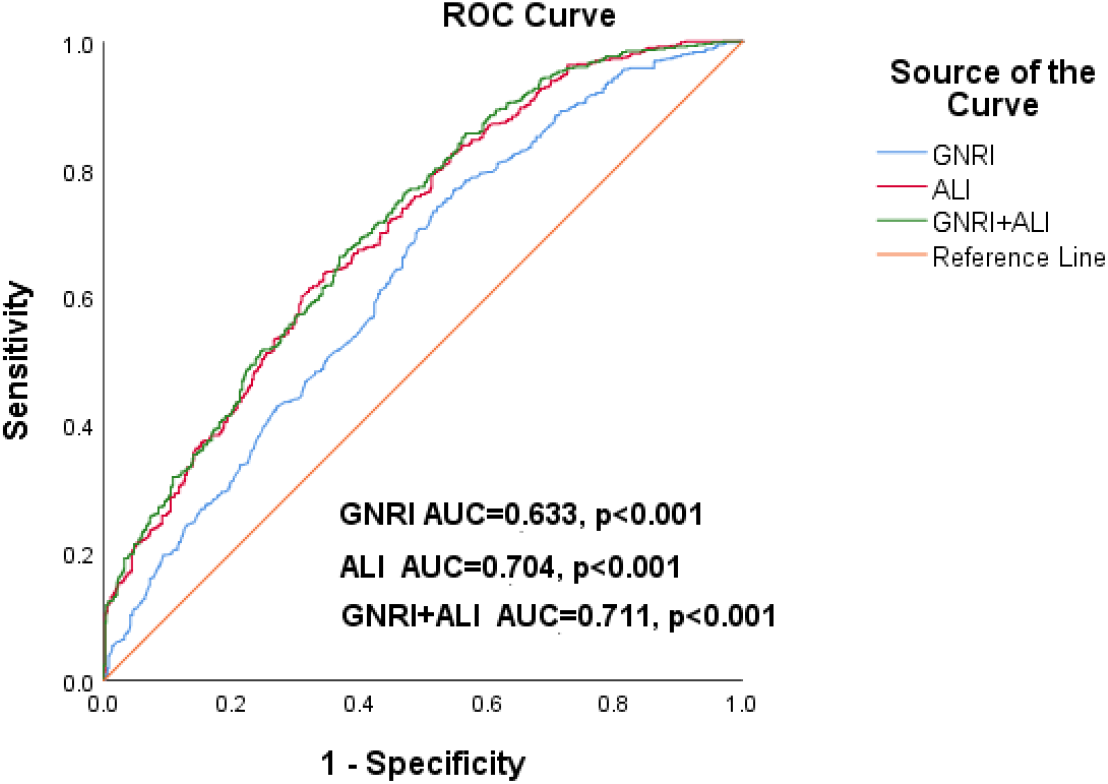
Time‐dependent receiver operating characteristic (ROC) curves of advanced lung cancer inflammation index (ALI), geriatric nutritional risk index (GNRI) and GNRI+ALI with reference line for all‐cause mortality in CHF patients. AUC, area under the curve.

**Figure 7.**
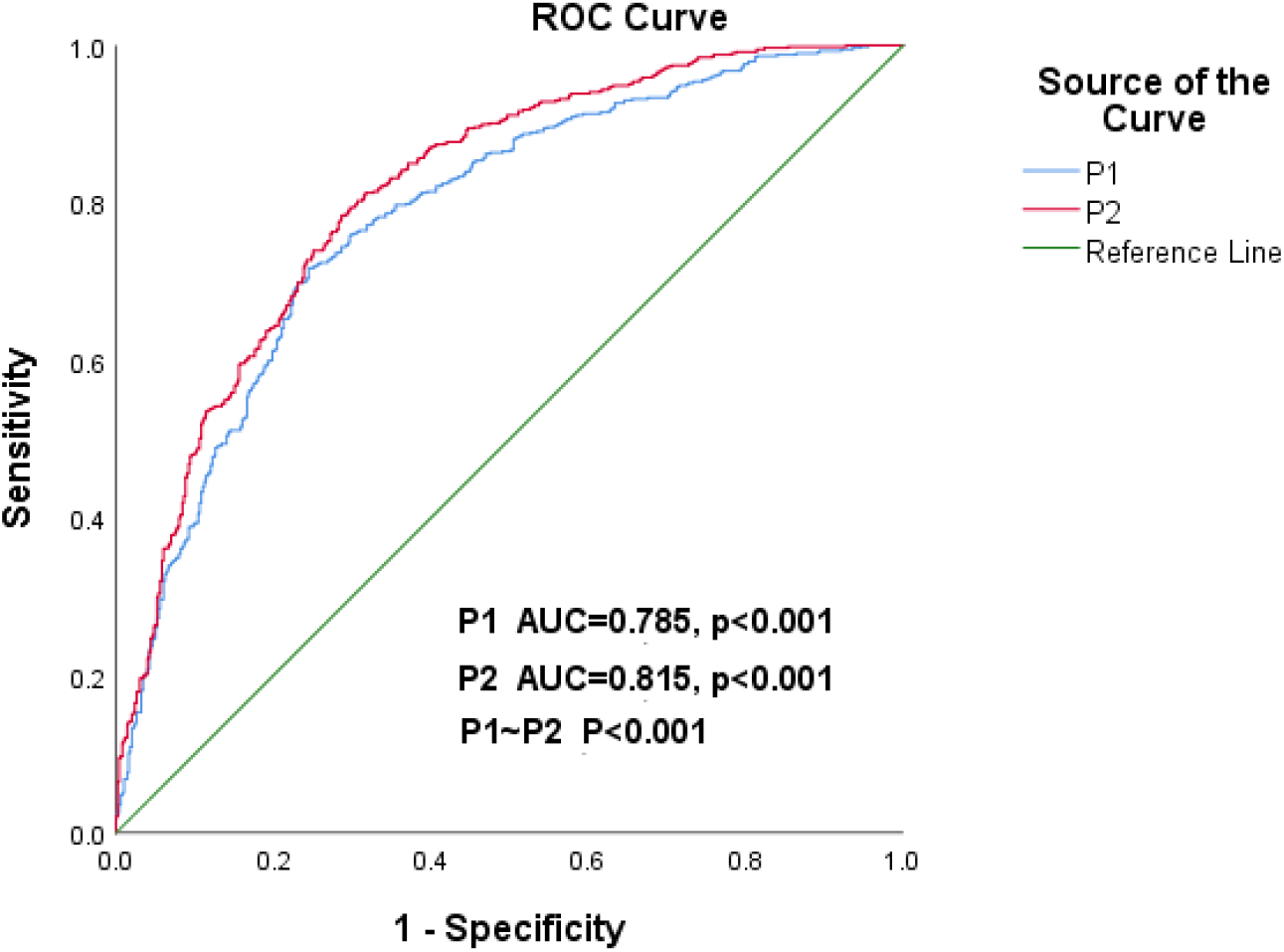
Time‐dependent receiver operating characteristic (ROC) curves of P1 and P2 with a reference line for all‐cause mortality in the HF patient. AUC, area under the curve. P1=age+NYHA+heart rate+systolic BP+diastolic BP+ALT+AST+creatinine+uric acid+eGFR+sodium +chlorine+lg BNP; P2=P1+GNRI+lg ALI.

## Discussion

This retrospective observational study was conducted to demonstrate that the ALI combined with the GNRI has a significant impact on the prognosis of patients with CHF. Multivariate Cox proportional hazards analysis revealed that both ALI and GNRI were independent predictors of all‐cause mortality in HF patients (ALI: HR 0.313, 95% CI 0.228‐0.430, p <0.001; GNRI: HR 0.966, 95% CI 0.953‐0.979, p <0.001). We divided the HF patients into Group 1 (ALI<24.60 and GNRI<94.41), Group 2 (ALI<24.60 and GNRI≥94.41), Group 3 (ALI≥24.60 and GNRI<94.41), and Group 4 (ALI≥24.60 and GNRI≥94.41) based on the median ALI and GNRI. The hazard ratios for mortality in the four groups showed that the risk of death was 3.113 times higher in Group 1 (p<0.001) and 2.067 times higher in Group 2 (p<0.001), with no difference between Groups 3 and 4 (p=0.307), using Group 4 as a reference. The time‐dependent ROC curves revealed that the AUC for the ALI combined with the GNRI was 0.711 (p<0.001), with a sensitivity of 67.34% and a specificity of 61.98%. The ALI combined with the GNRI has good predictive value for prognosis in CHF patients.

Factors contributing to CHF fall into four categories: first, common risk factors such as hypertension, ischaemic injury and metabolic syndrome; second, inherited heart disease such as hypertrophic cardiomyopathy; third, mechanical changes such as valve dysfunction; and finally, immune‐related causes including bacterial and viral infections and autoimmune reactions^13^. The calculation of ALI involves NLR, BMI and serum albumin, and GNRI is calculated from BMI and serum albumin. Of these, BMI and albumin are both common clinical markers that can reflect a patient’s nutritional status to some extent. There is a strong link between HF and malnutrition. Malnutrition can lead to hypoproteinaemia and a weakened immune system, which can aggravate HF and infections^14^. In contrast, as HF worsens, patients’ nutritional status deteriorates due to bowel oedema and reduced food intake, resulting in a vicious cycle^15–17^. However, obesity may also cause HF mainly because of the haemodynamic changes associated with activation of the renin‐angiotensin‐aldosterone system, enhanced sympathetic nervous system activity and mineralocorticoid receptor expression, and production of acute phase proteins and inflammatory cytokines^18^.

The ALI includes the neutrophil‐to‐lymphocyte ratio (NLR), so it is also an indicator of inflammation. Neutrophils activated by the immune response are complex cells capable of performing a large number of specialized functions. Experiments by Mantovani et al. have shown that overactivated neutrophils can lead to chronic inflammation and drive the expansion of innate and adaptive immune responses^19^. While neutrophils are important effectors of acute inflammation, neutrophil depletion in cardiac healing results in exacerbation of HF^20^. Lymphocytes are important immune cells involved in the pathogenesis of HF. T lymphocytes can regulate inflammation and related processes. These cells have been demonstrated to be involved in inflammation, hypertrophy, fibrosis and dysfunction of the heart^21^. Neutralization of T cells has been shown in animal models to have an attenuating effect on leukocyte recruitment and the development of cardiac hypertrophy. T‐cell depletion also reduces myocardial fibrosis, thereby modifying cardiac dysfunction^22^.

Our study combines ALI and GNRI to predict the prognosis of patients with CHF. The risk of death is highest when the ALI is <24.60 and GNRI <94.41, which reminds us of the need to pay clinical attention to the nutritional assessment of the patient and to improve the prognosis of the patient by regular nutritional therapy and by aggressive anti‐inflammatory treatment.

## Conclusion

Our study shows that the ALI and GNRI are independent predictors of prognosis in patients with CHF. There was a mild positive linear correlation between the ALI and GNRI, and there was no interaction. The risk of all‐cause mortality for CHF patients from highest to lowest was as follows: Group 1 (ALI<24.60 and GNRI<94.41)>Group 2 (ALI<24.60 and GNRI≥94.41)>Group 3 (ALI≥24.60 and GNRI<94.41=Group 4 (ALI≥24.60 and GNRI≥94.41). ALI and GNRI have a significant impact on the prognosis of CHF patients, and the ALI combined with the GNRI can be a good predictor of the prognosis of CHF patients.

## Data Availability

All the data involved in the manuscript are available.

## Limitations

This study is a retrospective observational study and may have some degree of selection bias. Further prospective studies are needed to test the prognostic impact of the ALI combined with the GNRI in patients with chronic heart failure. Our primary study was in patients with NYHA class III or IV. Therefore, some study results may not be available for patients with comparatively mild HF symptoms.

## Sources of Funding

The study was funded by the Yunnan Provincial Health Commission Clinical Medical Center (ZX2019‐03‐01) and by the Applied Basic Research Program of the Science and Technology Hall of Yunnan Province and Kunming Medical University (Project No. 202301AY070001‐130).

## Disclosures

None.

## Notes

### Competing Interest Statement

The authors have declared no competing interest.

